# EffiCare: Better Prognostic Models via Resource-Efficient Health Embeddings

**DOI:** 10.1101/2020.07.21.20157610

**Authors:** Nils Rethmeier, Necip Oguz Serbetci, Sebastian Möller, Roland Roller

## Abstract

Recent medical prognostic models adapted from high data-resource fields like language processing have quickly grown in complexity and size. However, since medical data typically constitute low data-resource settings, performances on tasks like clinical prediction did not improve expectedly. Instead of following this trend of using complex neural models in combination with small, pre-selected feature sets, we propose `EffiCare`, which focuses on minimizing hospital resource requirements for assistive clinical prediction models. First, by embedding medical events, we eliminate manual domain feature-engineering and increase the amount of learning data. Second, we use small, but data-efficient models, that compute faster and are easier to interpret. We evaluate our approach on four clinical prediction tasks and achieve substantial performance improvements over highly resource-demanding state-of-the-art methods. Finally, to evaluate our model beyond score improvements, we apply explainability and interpretability methods to analyze the decisions of our model and whether it uses data sources and parameters efficiently.^1^

## Introduction

For many machine learning tasks we can observe that more complex models with more data tend to outperform the previous state-of-art model. Particularly deep learning approaches can implicitly learn to extract and apply task relevant features^1–8^. However, complex models require more compute resources and large amounts of data to learn well – usually the more data the better. In clinical contexts this might be an issue as large datasets are often not available. For example, on the 2017 four-task clinical prediction benchmark by Harutyunyan^92^, the paradigm of applying complex models taken from other fields, but limiting training data to a small set of expert-selected features has resulted in a stagnation in performance – see Table 2 †. Limiting the amount of input features is based on two common assumptions. From the medical perspective, limiting features to known ones increases trust and model interpretability. From a machine learning perspective, it is common to limit the number of input variables (features) and discard rare ones, in an effort to make learning easier. A welcome side effect of such limitation is that memory, compute and manual labor requirements are minimized to meet real world time and cost limitations. However, feature limitation by expert bias prohibits the discovery of new and unforseen correlations. Moreover, a highly limited feature set combined with relatively small dataset does not provide enough input information to fully utilize deep learning methods. This problem is compounded by an increasing trend to adopt the latest large, complex models with many learnable parameters from domains like computer vision or natural language processing^1–8^, where data types are much more homogeneous and large scale data can be readily exploited. On the other hand, Tomašev et al.^10^ provide a recent example of the performance benefits gained from using all instead of an expert-selected feature set. However, when using hundreds of more features with complex models, the resulting learning setup quickly becomes impractical for hospital deployment, due to the large memory, compute and complex model interpretability. For low-data tasks we can use small embedding-pooling based models that were originally designed as text classifiers. Those small models outperformed complex, 29-layered, convolutional networks, while using orders of magnitude less parameters and compute^11^.

By combining and extenting upon these insights from other fields we propose and demonstrate the following ‘less-but-better’ approach of using large data, with small models, called `EffiCare`. We *introduce a resource-efficient feature-embedding method*, along with a *lightweight neural architecture* to process electronic health records (EHR) efficiently. This allows our approach to be able to encode and process a large variety of information performantly. We test our method on four different clinical prediction tasks found in the context of intensive care unit (ICU)^9^. Using smaller resource-efficient models with feature-embeddings, we can handle an amount of data that would be resource-prohibitive for large models – i.e. we train over hundreds of features, while previous (complex) models used only a few hand-picked features^5–9^, given comparable compute setups. By combining two unsupervised pre-training methods^11,12^, we simplify recent ideas of embedding patient health event histories as *time-stamped embedding sequences*^5,7, 8^, to ‘embed-away’ the sparsity of health records. The proposed embedding and model approach greatly reduces the manual labor required to add data sources and test new models, which enables faster development iterations. As a result of fast development we quickly realized that less complex, pooling based models greatly outperform complex recurrent, convolutional or self-attention approaches – both in terms of task scores and resource efficiency.

Finally, we focus on model trust and transparency through ‘model understanding’^13^, i.e. interpretability methods^14,15^, to inspect the model, and model ‘decision understanding’^13^, i.e. explainability^16^, to identify important medical events for predictions. Combining ‘model and decision understanding’ allows us to verify that our models not only optimize task predictions, but also implicitly learn to identify important biomarkers for each prediction task and that they use data sources and model components efficiently, i.e. with little redundancies. Overall, we improve performance and minimize labor, time, data and hardware use to help remove these major obstacles in applying state-of-the-art prognostic models to ease the day-to-day hospital application.

### Benchmark Dataset

MIMIC-III^17^, is an anonymized public database containing electronic-health-records (EHR) for over 40,000 patients from intensive care units (ICU). The data is longitudinal, heterogeneous and irregularly sampled. Furthermore, there are duplicate entries and erroneous input by the medical staff. For evaluation, we use benchmark tasks provided by Harutyunyan et al.^9^. We refer to this work as *benchmark* or *Haru17/19. The benchmark includes four different tasks, namely *In-Hospital Mortality, Decompensation, Length of Stay* and *Phenotyping. In-Hospital Mortality* task predicts whether a patient will die during their stay at the hospital based on the first 48 hours of ICU admission. *Decompensation* task predicts at every hour whether the patient will die in the next 24 hours. *Length of Stay* is the task of predicting the remaining number of hours of a patient in ICU at each hour of stay. *Phenotyping* addresses the task of classifying the diagnosis (multilabel out of 25 acute care conditions) at the end of the patient’s stay. Each task uses the data of an individual ICU stay up to the prediction time. The resulting benchmark dataset consists of 33,798 unique patients with a total of 41,902 ICU stays and over 250 million clinical events. Each sample corresponds to an individual ICU stay of a patient. Our experiments use the same cohorts, including the same training, development and test data splits. To limit information leakage, the splits are made based on individual patients, not ICU stays.

### Embedding Sequential Data

Most related work in the context of prognostic deep learning models use a set of fixed features which are inserted into a sequential neural architecture^6,8–10^, expecting each particular feature at a fixed position. Opposed to that we would like to avoid a fixed set of predefined features or techniques such as imputation. In natural language processing we can technically use an unlimited amount of sequential embedding features for words to feed word co-occurence semantics into a machine learning model. Such embeddings are multi-dimensional vectors, that are pre-trained using self-supervision^11,12^ such that words that share common contexts are placed close to one another in the vector space, i.e. have a smaller cosine similarity. Inspired by this idea we encode diverse sets of events and their non-scalar values as vectors as described in the following. To gather events, we use the same source data tables from MIMIC-III as the benchmark. The benchmark paper constructs a set of 17 hand-picked features, which they pre-process by merging duplicate events and cleaning values. We instead use all 4336 events from those tables. Namely, `CHARTEVENTS` containing the electronic chart that displays patient’s information relevant to care, `LABEVENTS` containing all the laboratory test results and `OUTPUTEVENTS` consisting any output fluid excreted by or extracted from the patient, such as urine output or drain after surgery. In the training set, patients have 87 hour stays on average, where 92% of these hours have events in `CHARTEVENTS`, 15% in `LABEVENTS` and 12% in `OUTPUTEVENTS` tables. If an hour has events, it has on average 50 in `CHARTEVENTS`, 16 in `LABEVENTS` and 1 in `OUTPUTEVENTS` tables. The same event, e.g. heart rate measurement, can occur multiple times within an hour. Each event consists of hours elapsed since admission, a label such as Heart-rate or Heart-rhythm, and a corresponding value such as 89 or “asystole”.

To embed the event label into a vector representation we use *randomly initialized embedding vectors* and pretrained *FastText embeddings*. To obtain pretrained FastText embeddings, event labels occurring together within a time step are concatenated and written into a file – see Figure 1. Using this sequence of feature labels any word embedding method can be applied. We use the FastText^11^ skip-gram method to learn those embeddings because FastText incorporates sub-word information in token embeddings. In this way, labels that share sub-words are close-by in the embedding space, e.g. heart-rate and heart-rhythm. Learning embeddings for sub-words also helps us mitigate problems of changing the distribution of tokens by decreasing the number of categorical features by splitting each into many with concatenated values. After obtaining these embeddings, we use them as a non-trainable, or frozen, embedding look-up table in our models. That way we can feed an event sequence into this table, which turns them into an embedding sequence. Note, values in MIMIC-III are highly heterogenuous and unclean, e.g. 89, “SR (Sinus Rhythm)”, “1cm”, “24/24”. To handle such input we first obtain unique feature labels by concatenating event names and any value that is not a scalar, e.g. judgement-intact, ventilation rate-24/24. After obtaining the vector representation for the event label, scalar feature value and time stamp we concatenate these features into a single ‘concept-value-time’ embedding vector – Figure 1. Scalar feature values are discritized and encoded with a one-hot vector. For each individual feature we calculate uniform buckets for values using the training set. Each bucket represents a value range of that feature. In addition, two further buckets are used to deal with outliers. Categorical feature values are already included in feature label embedding and are represented as zero vectors here. Furthermore, time bucket *t* in hours is included both in logarithmic and exponential form: log(*t* + 1), exp(*t/*1000)*−* 1. Using this calculation normalizes the hour format (which can go up to 1500 hours) in two non-linear ways^18^. The complete pipeline is shown in Figure 1.

**Figure 1:**
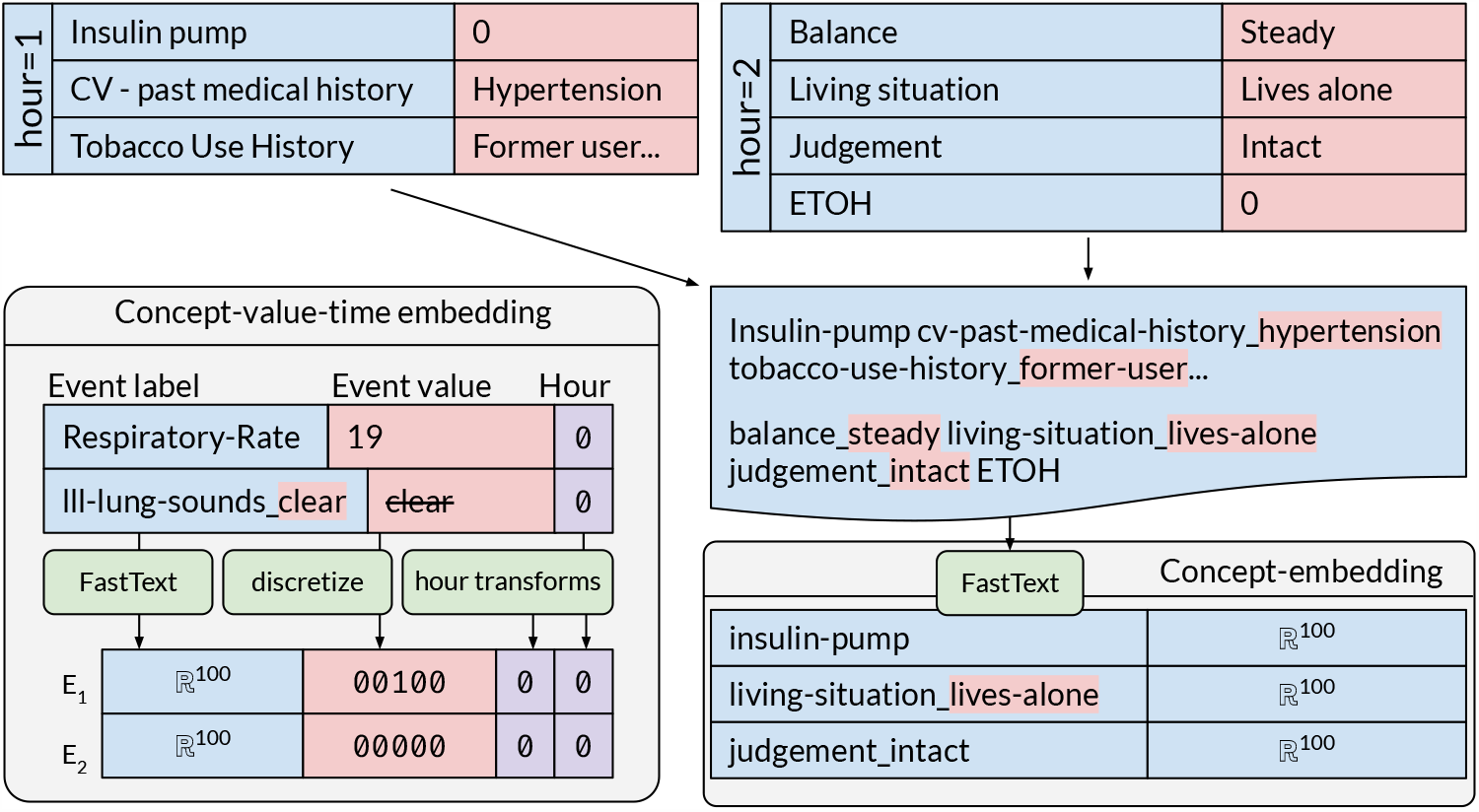
**Concept-embeddings:** Feature labels in an hour slot co-occur as a sequence of concepts, to train concept-embeddings via FastText – see upper and right image portion. **Concept-value-time embeddings**: Concept-embedding are concatenated with binned feature values and encoded time stamps. For example, the value 19 of label *Respiratory_Rate* is discritized as 00100, assuming that 19 is a value in the middle range of *Respiratory_Rate* based on training set values. Categorical label-values like *clear* for *lll-lung-sounds* are assigned a zero vector 00000.

### Prognostic Multitask Model

To efficiently integrate the large amount of data features occurring in the ICU we use a small model with as few parameters as empirically necessary. This enables us to train the model within a reasonable amount of time, despite using thousands of features – i.e. our best performing model trained in 40 minutes*×* 14 epochs on a single GPU^3^. During early experiments we also tested deep and shallow GRU-based and CNN based architectures, which were significantly slower to train but performed worse than the small time-pooling based model. Figure 2 provides an overview of our prognostic multitask model. The events from different sources (e.g. chart events) are embedded and collected within ‘1-hour-buckets’ containing all embeddings occurring within a single hour *t* for that source. The size of each bucket is patient-dependent according to the maximum number of features occurring within an hour for that source. For hour buckets with few features we zero pad. Each embedding *e*_*t*_ from bucket at hour *t* is fed into a fully connected layer (FC) with ReLU activation, followed by max, avg and normalized sum pooling over the events. The pooled outputs are concatenated to obtain an hourly patient embedding *s*_*t*_, combined with the pooled outputs of the other event sources. This step (A) is applied for all hours of all events since admission (until the current prediction time *s*_*now*_) to generate a sequence of hourly patient embeddings from *s*0 to *s*_*now*_. Finally, all patient embeddings up until prediction time are pooled over time. The pooled outputs are then concatenated with the demographic embedding (B), into a patient embedding *p*_*now*_ that aggregates the patient’s state from admission up until hour *now*. This patient embedding then feeds five fully connected layers for the four prediction tasks (see right side of figure). As in Harutyunyan et al.^9^, length of stay is predicted as both 10-way classification *FC*_*LC*_ and regression *FC*_*LR*_. Task losses *L*_*task*_ are weighted: .1*L*_*LC*_ + 1*L*_*LR*_ + 2*L*_*DECOMP*_ + .2*L*_*IHM*_ + 1*L*_*P HENO*_. While max and average pooling are commonly used normalized sum pooling, see equation 1, was introduced by^19^, to account for the accumulation of risk through time. In contrast to average pooling, this sum pooling also ignores zero-padding, resulting in an additional implicit learning feature.

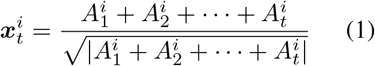

**Figure 2:**
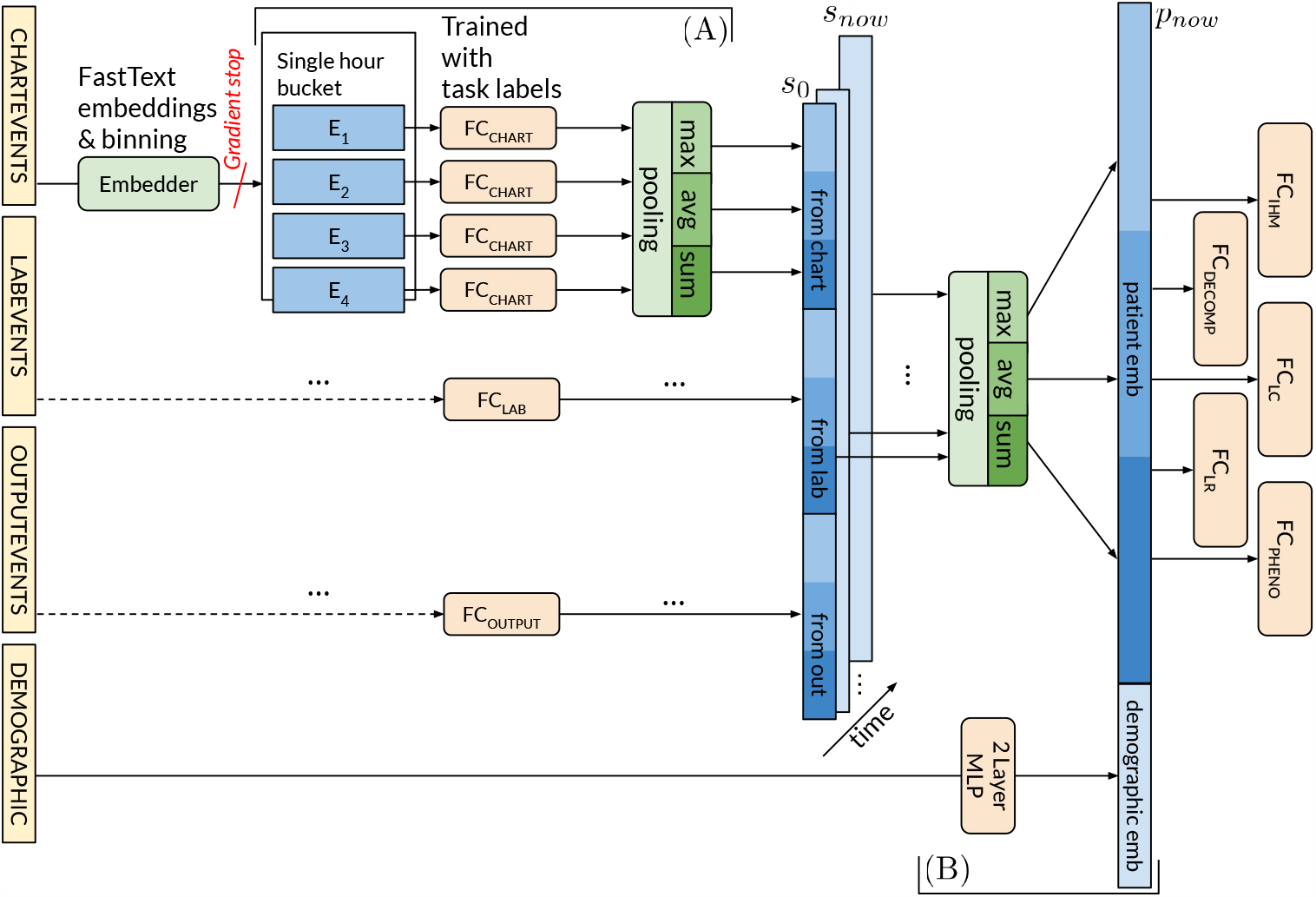
Our event-time-pooling model: Green boxes are modules without or with fixed parameters (Embedder). The left model half shows how a single time-step of event embeddings is pooled per data input source (4 yellow sources). We then pool over all time steps and data source features to discard unhelpful information.

### Configuration and Experimental Setup

For the following experiments, we trained feature representations using a FastText with a window size of 15 and default parameters with 100 dimensional embeddings. Scalar values are discritized into 10 uniform bins with two additional outlier bins for the 0.01 and 0.99 percentile values. Bins are calculated over the training set and are fixed at test time. This results in a one-hot vector of length 12 for scalar-valued features and a zero vector for any categorical feature. The fully connected layer for each table has 50 dimensions. The demographic input is sent through a two layer MLP with dimensions 40 and 20 and ReLU activations in-between. Each task is predicted by a corresponding fully connected layer using this representation.

We train with mini-batch size 16. For regularization, we apply input dropout with probabilities 0.15 for events and 0.1 for whole time-steps. The demographics network is regularized with 0.3 dropout. All tasks, but especially decompensation, have imbalanced label distributions. Thus, we use class weighting while calculating the training loss. This is equivalent to over-sampling patients that correspond to less frequent labels. For the multitask loss, we use the same weighting as in the benchmark paper^9^, except for the fact that we train regression and classification for length of stay at the same time: 0.2 for in hospital mortality, 2 for decompensation, 1 for length of stay classification, 0.1 for length of stay regression and 1 for phenotyping. As optimizer we use Adam with Pytorch defaults, i.e. learning rate 0.001 and betas 0.9, 0.999. Since a few patient histories are disproportionately long, we consider only the first 30 days of each patient’s stay at the hospital during training, as done by^7,8^. At test time, we predict over all time-steps. For development and model selection, we used the training and validation set defined by^9^. The test set from^9^ is used only to report results.

## Results

See results in Table 2. As ROC scores overestimate performance under class imbalance^20^ we only discuss the results related to AUC-PR and Kappa. In case of *Phenotyping* previous works do not provide AUC-PR results. While a higher score represents an increase in performance for most evaluation measures, in case of MAD, a lower score is better.

### Sequence embeddings of patient features majorly boosts performance

First, we evaluate our embedding-based methods (†) against the previously best single and multitask models (*)^6–9^ in Table 2. For each baseline work(*), we list the best model results only. The *Haru17/19 benchmark has the oldest results, originally published in 2017, then republished in a Nature 2019 issue^9^. The first two works (*) only use the 17 features selected by *Haru17/19. Notably, more recent complex models by Ma and Song *did not beat the less complex baselines* by *Haru17/19. We also see that multitask learning performs better. These observations suggests that we should use less complex, multitask learning models with more than 17 training features. Our approaches, that use *all 10,461 benchmark feature embeddings* with our comparatively simple multitask models(†), outperform the base-lines (*) on all tasks by large margins, where some scores such as length of stay MAD and decompensation AUC-PR almost double. Furthermore, as Table 1 shows, even though our approach uses many more features, it has a much smaller number of trainable parameters. Thus, small neural architectures that are designed for low-resource scenarios by using fewer parameters are not only easier and faster to train, but also boost performance.

**Table 1:** Number of trainable parameters and features. Each categorical event-value counts a as a single feature. For comparison, the 17 features used in *Haru17/19 result in 64 individual features. *FastText only 17* counts non-preprocessed events used to construct the 64 features from *Haru17/19^9^.

| Model | # Parameters | # Features |
| --- | --- | --- |
| *Haru17/19 MC-model (MT)<sup>9</sup> | 1,225,253 | 64 |
| FastText embeddings | 70,528 | 10461 |
| FastText event count > 100 | 70,528 | 6165 |
| FastText only 17 *Haru17/19 | 70,528 | 75 |

Further, we can conclude that *using all available features drastically benefits learning* and that multiple tasks may benefit each other through shared, pretrained event embeddings (see Embedder in Figure 2). Note that, random embeddings do not require embedding pretraining, i.e. requires the least modeling effort. However, when we exploit embedding pre-training via FastText^11^ we improve on all metrics except length of stay Kappa.

Finally, as an in-practice near zero-effort optimization, we retro-fit our event embeddings using the autoencoding method by *Reth19^124^. We select optimal *Reth19 embeddings using the validation set, to gain a 4.1 point better decompensation test score without using extra labor or hardware resources. For completeness, we also mention three negative (dead-end) insights. We initially tested many more complex models, but since using CNN and GRU layers strongly underperformed in prediction and compute performance compared to our smaller models, we choose to not pursue complex architectures. Further, excluding the number of hours elapsed since admission in the input embeddings hurt performance moderately, while using only max pooling considerably hurt performance. Telling us that average and sum pooling are important components.

### Early expert bias for feature limitation hurts performance

Next we analyze if early domain expert-based bias, like feature selection or removing rare features, impacts model performance. As done in the benchmark paper^9^, we use only the 17 hand-picked features to train our model, except we do not do any feature clean-up (see *FastText only 17 *Haru17/19*). Table 2 shows a large drop in overall performance compared to the all-features (†) *FastText embeddings* model, except for *Length of Stay* Kappa. However, even in this low-data setting, our approach still performs much better or similar to much larger state-of-the-art models, indicating that small models may be better suited to the data setup. When not training with infrequent features († FastText event count *>* 100), decompensation task performance decreases. Hence, relevant rare features were dropped. Thus, when delaying domain biases like *expert feature selection* or rare event filtering until *after model training* we can greatly improve test set prediction performance or identify resource reduction relevant factors. Results for removal of demographic features († FastText w/o demographics) are described in the interpretability-detail section (next page). NOTE: *Standard deviations* or small fixes can be found in the repo.

**Table 2:**
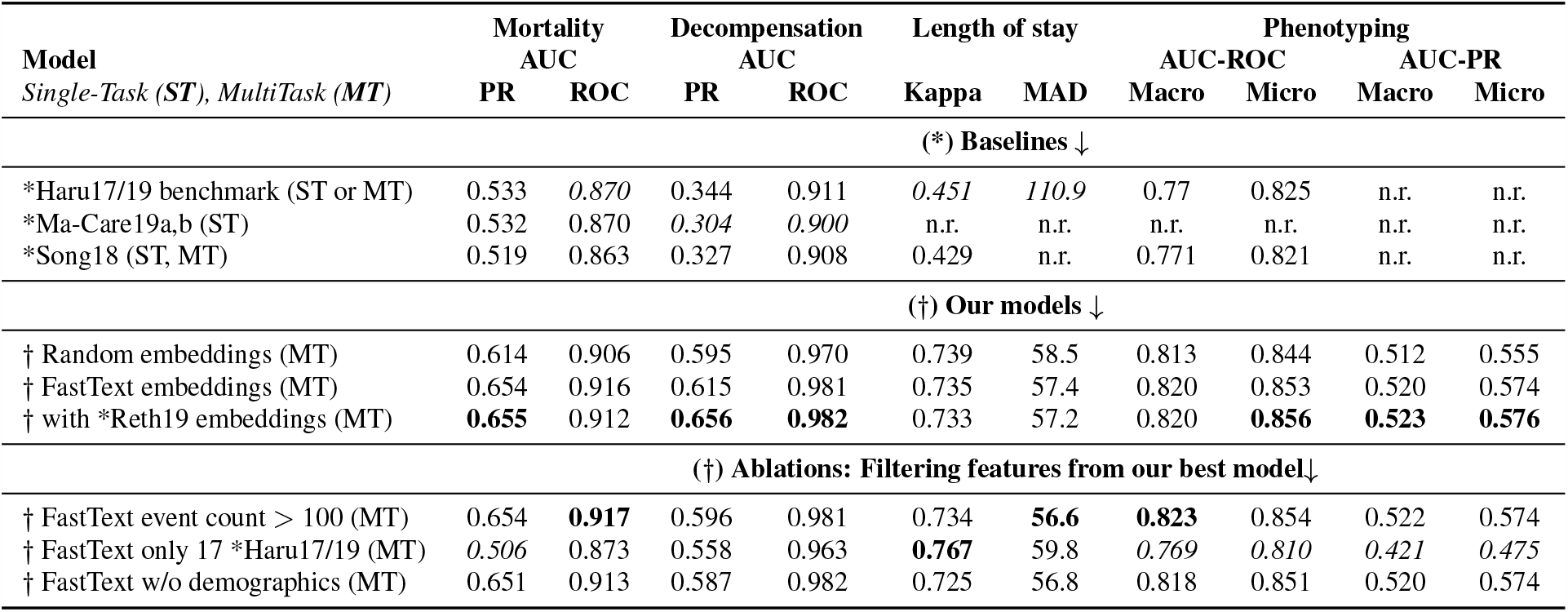
**\*Haru17/19 Benchmark test set performances for the SOTA models (*) vs. our multitask, all features model** († `EffiCare`). **\*Haru17/19** results use only 17 expert-selected features with LSTMs^9^. *Ma-Care19a,b are baseline results from^7,8^ (Dilated CNN, Transformers), while *Song18 is from^6^ (Transformers). **\*Reth19** are FastText embeddings retro-fit via^12^. **Best**, *worst* and n.r. (not reported) scores. (ST, MT) indicate single and multitask models.

### Interpretability for ‘model understanding’

Automation bias, is an *over-reliance on decision making technology due to high system complexity and low understanding*. Thus, as suggested by^13^ we use ‘model understanding’ techniques to peer into our models’ black-box to increase awareness of its capabilities and limitations by inspecting learned abstractions. Specifically, we analyze how our best model combines and filters patient features and how important different model components are overall for predicting each task. Using intepretability by^14^, we allow ‘model understanding’ at both a detailed and an overview level.

First, we gain a *detail-view* of how a model represents domain knowledge from chart or lab events for all tasks by adapting the interpretability concept of “aggregating and visualizing what features the neurons in a model prefer” from^14,15^. This concept was introduced in image recognition to visualize a model’s learned abstractions like eyes or noses^15^. The method was recently extended to visualize language model word feature abstractions^14^, which we adapted to visualize what events our model components prefer during. Secondly, we aggregate activations of the learned patient representations (*patient emb* in Figure 2) to gain an *overview* of how important different types of either domain knowledge (data sources) or model components are for predicting each of the four tasks.

### Interpretability detail-view, reveals (non-)redundancies

To visualize what low-level filters learn about individual data sources like chart or lab events, we collect the “preferred (maximally activated) features” of each neuron in *FC*_*CHART*_ and *FC*_*LAB*_ over the training set. Thus, each ‘filter neuron’ in *FC*_*CHART*_ or *FC*_*LAB*_ forms a distribution of ‘preferred features’ using the method and implementation^5^ proposed by Rethmeier et al.^14^, who similarly visualized knowledge abstraction during unsupervised language model training. In Figure 3, we see which patient features are the most active (preferred) in two neurons of our model’s learned low-level feature filters. Note that, we see one chart neuron plus one lab neuron out of a total 150 filters – i.e. 50 filters per chart, lab and body-output events.

**Figure 3:**
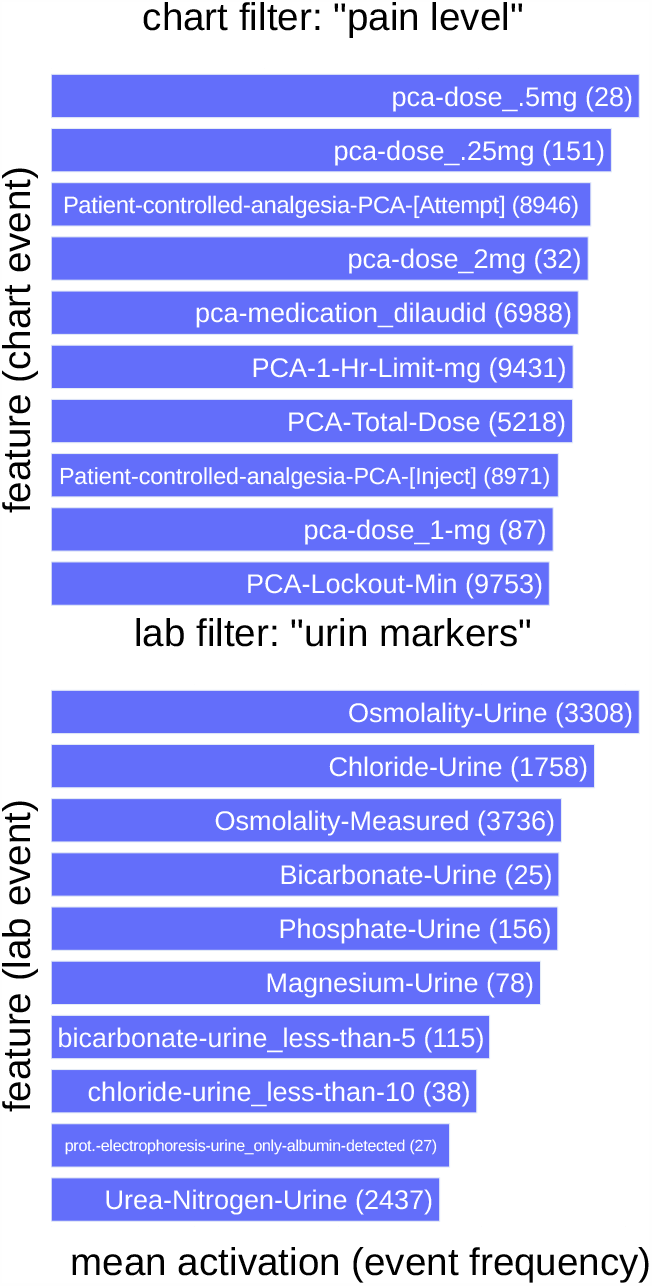
Detailed interpretability view: Top-10 “preferred features” of two patient-event level filters. Two filters *H*_*i*_ from *FC*_*CHART*_ and *FC*_*LAB*_ (Figure 2). The chart filter abstracts pain management events, the lab filter captures urine indicators. **(Rare features)** like ‘Bicarbonate-Urine’ matter – see **(feature count)**.

The presented *chart data filter* shows a strong activation-preference on information regarding the need for self-controlled pain-relieve – i.e. the patient’s perceived pain level. The presented *lab filter* is most active for features that involve urine lab values. Thus, these filters *implicitly learned to abstract and cluster knowledge* about pain and urine related health indicators, even though *we did not model any explicit feature clustering or preprocessing* as done in older, manual feature-engineering approaches. When looking at the other filters, we found that many filters learned to focus on specific clinical contexts: bed-laying pressure points, cardiology-cholesterol measures, blood health, self-sufficiency, and diet.

Moreover, we found that data sources, e.g. body-output data, that have fewer features than others, have an increasing number of filters with empty feature preference distributions. This suggests that those filters may not be necessary and could be removed to shrink the model and therefore its hardware requirements. Interestingly, we also observed that non-empty filters form unique feature clusters – i.e. no duplicate clusters. This indicates that these filters learned to *avoid abstraction redundancies*, or as representation learning terminology puts it, our model learned disentangled representations^6^. Practically, efficient data representation results in a model that uses computations and memory in terms of its parameters efficiently. Finally, we noticed that chart event filters such as those seen in Figure 3 already encode demographics such as sex and weight. Accordingly, after removing the demographic input data and its 2 layer MLP component, test set performances were nearly unaffected – see *† FastText w/o demographics (MT)* in Table 2. Thus we conclude that, *interpretabilty helps identify and then remove unnecessary (redundant) model components and data sources*. Importantly, such ‘model understanding’ only visualizes model abstractions, not how these abstractions are weighted to solve tasks (‘decision understanding’), as we will do below.

### Interpretability overview, data or model components relevance per task

To gain an overview on how each prognostic task uses features from chart, lab and output events we collect learned patient embedding representations (see Figure 2) for all patient histories in the development set. Next, we multiply the representations with each of the four classifier weights. In this way we can investigate how strongly (active) each task weights specific data sources and pooling information. In Figure 4 we see that neither of the two tasks uses body-output feature information and that data and pooling importance vary per task. However, when we remember (see data section) that only 12% of patient-buckets have `OUTPUTEVENTS`, while 92% and 15% have `CHARTEVENTS` or `LABEVENTS`, we understand that output events still matter, and that lab events are very important^7^, even though chart events dominate by raw frequency and hence weighted activation. For decompensation sum and average pooling over a single time step (event-level) cause both strong positive and strong negative impacts on the prediction score. Also, chart events are the most important here. At event-level, average and sum pooling have strong impacts on predictions, while at the pooling-over-time-level, we see that max pooling is the most impactful. We also see that average pooling over time causes only positive impacts. Thus, each task uses data sources and network components differently.

**Figure 4:**
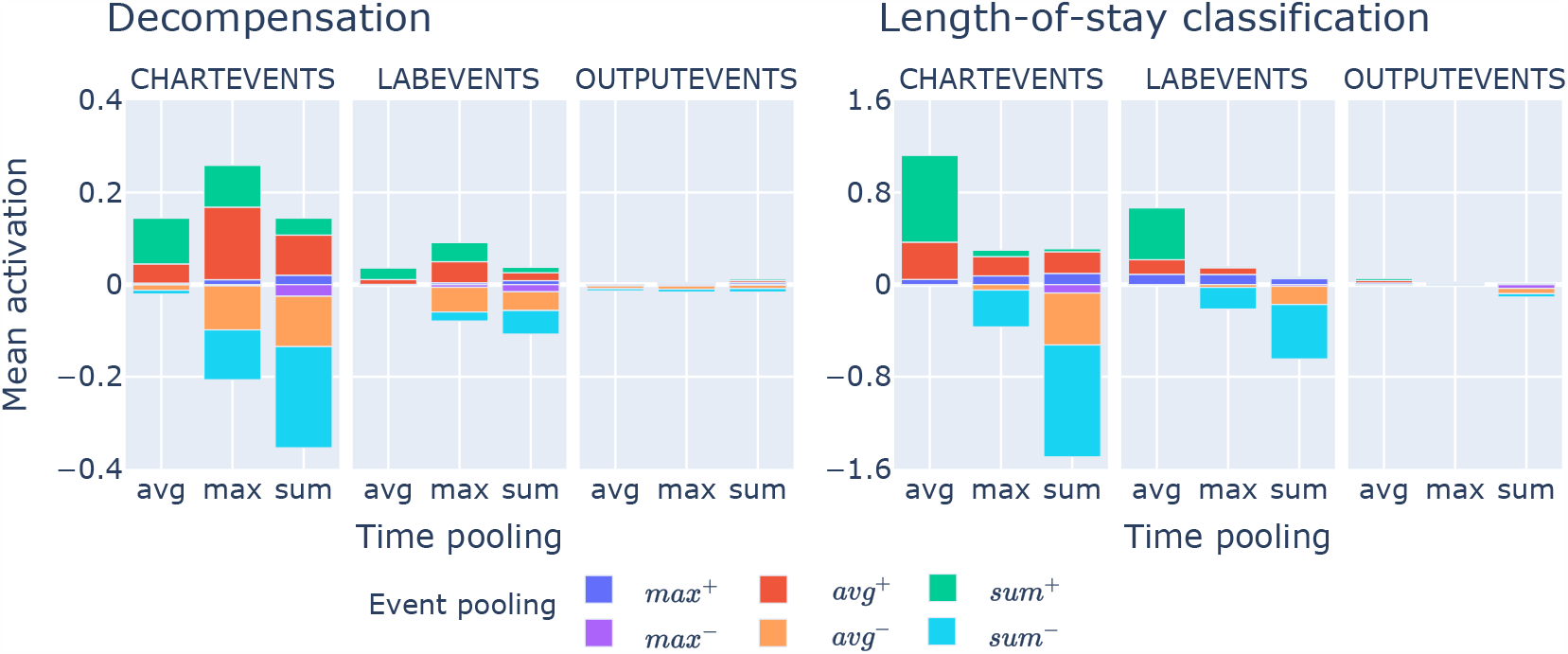
Feature type and pooling mechanism relevances for two tasks. The x-axis shows relevances for max, average and sum pooling over time (high level patient embeddings). The color bars express relevances of pooling *over events of a single time step* (low level data filters *FC*_*CHART*_, *FC*_*LAB*_ and *FC*_*OUT P*_) – see Figure 2.

### Explainability for per-patient ‘decision understanding’

So far we only looked at understanding how the model abstracts data and uses its component to model the four tasks. However, to also provide ‘decision understanding’^13^ of what features are most relevant or impactful for a specific prediction, we use the explainability method of integrated gradients (see^13^). We again split our analysis into overview and detail. This allows us to see how (counter-)indicative specific event value combinations are in for predicting patient mortality overall compared to how important events are in a single patients history.

### Explainability overview of the most and least predictive features

Using the popular integrated gradients method^16^, we calculate for each development set feature its impact on the prediction score of true positive in-hospitality-mortality predictions. Table 3 shows that features like *heart-rhythm=asystole* or *Gastric-Emesis* have a strong positive influence, whereas *Drain-Out-#1-Tap* or *Ultrafiltrate-Ultrafiltrate* have a strong negative influence on a positive prediction. “Other” describes four identically labeled hematology events from `LABEVENTS` that are related to fluids: Cerebrospinal Fluid, Joint fluid, Pleural, Ascites. As we expected, high impact features describe events that likely occur close to an in-hospital-mortality. Importantly, we see that some infrequent features have a strong positive or negative influence on the classification, meaning they should not be prematurely filtered.

**Table 3:** Explainability-overview for In-hospital-mortality: Data features (events) that raise or lower correct predictions for the in-hospitality-mortality task. Left: 7 most raising features. Right: 7 most score-lowering features. * Infrequent features are important for the task.

| top mortality indicating features | ↑ impact | frequency | top counter-indicative features | ↑ impact | frequency |
| --- | --- | --- | --- | --- | --- |
| code-status=comfort-measures-only | 0.87 | 161 | Drain-Out-#1-Tap* | -0.89 | 95 |
| heart-rhythm=asystole | 0.82 | 194 | Jackson-Pratt-#1 | -0.81 | 4102 |
| Gastric=Emesis | 0.79 | 1174 | Ultrafiltrate-Ultrafiltrate | -0.67 | 4163 |
| Other* | 0.74 | 119 | Drain-Out-#1-Sump* | -0.63 | 21 |
| code-status=cpr-not-indicate | 0.68 | 1150 | sputum-source=expectorated | -0.61 | 9951 |
| code-status=comfort-measures | 0.66 | 3044 | Drain-Out-#2-Other* | -0.54 | 140 |
| code-status=do-not-resuscita | 0.66 | 50477 | diet-type=diabetic | -0.54 | 14081 |

### Explainability detail: (counter-)predictive feature in a patient’s history

Figure 5 shows the attributions of input events to a correct in-hospital mortality prediction of a *single patient*. The prediction is done at 48th hour as per the task definition and the input has 3438 events in total. The figure shows events with a high (*>* 0.2) positive or negative impact (attribution) on the mortality of patients that eventually died by hour 368. The events from the initial 48 hours that are most indicative for the death of the patient are *Foley=3, abdominal-assessment=ascites, Urea-Nitrogen-Urine=3, patient-location=cc6b Lactate=4 and Anion-Gap=4*. Most counter-indicative are features like *Sputum-source=expectorated, Urea-Nitrogen=2, Lactate=2 and Foley=3*. This visualization lets us analyze which events are most critical to a patients condition over time, according to the trained model.

**Figure 5:**
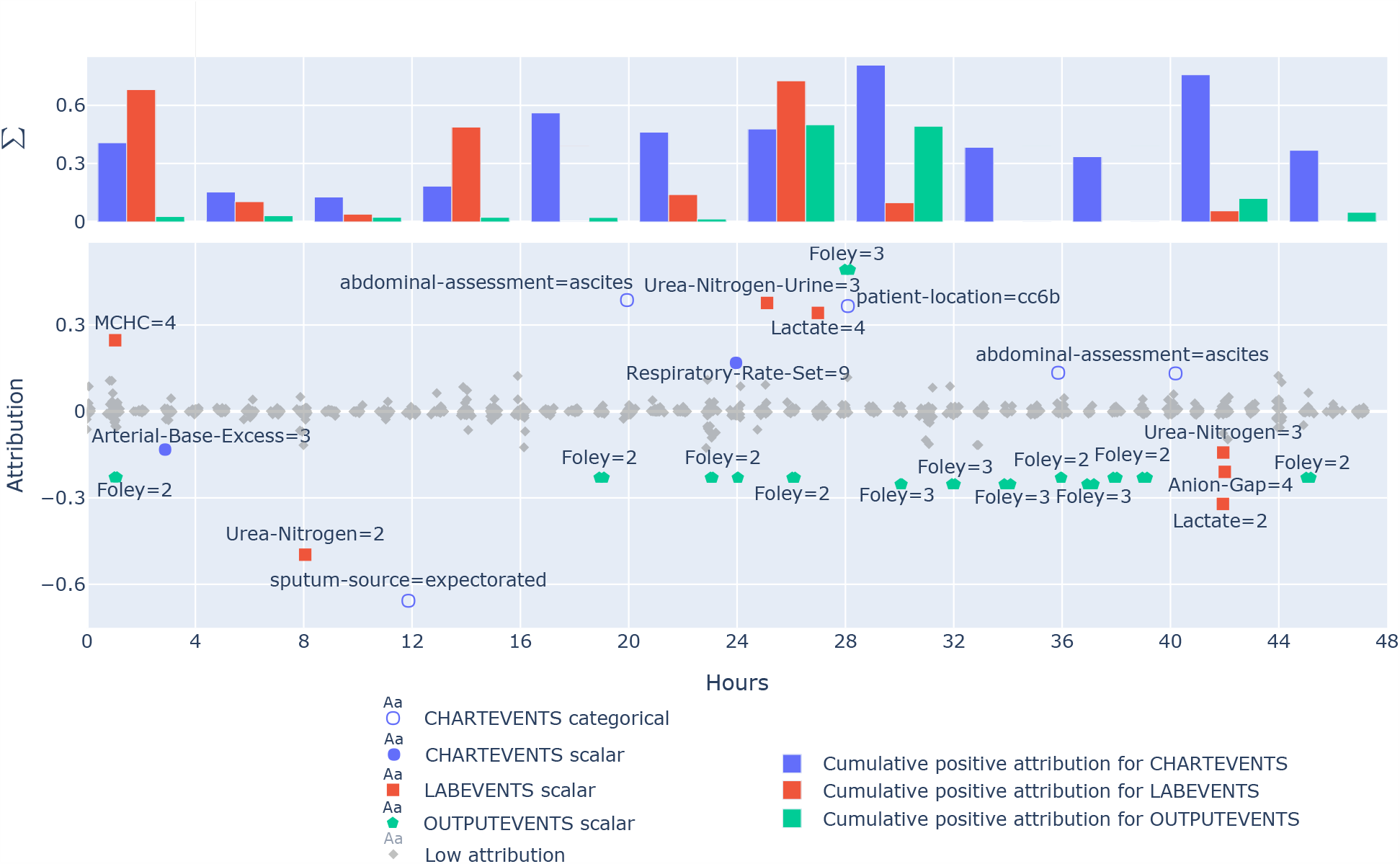
A single patients 48 hours integrated gradient explanation for a correctly predicted in-hospital mortality. Equals are categorical values, or the bin of a scalar value, where 1 is the lowest and 10 the highest bin (Outlier bins 0 and 11 are infrequent.) The bars show accumulative positive attributions for events in 4 hour blocks for different data sources. Colored events have a mortality prediction attribution (*>* 0.2). Negative attributions are counter-indicative.

## Discussion & Conclusion

We presented a novel, resource-efficient patient event sequence embedding method and model, called `EffiCare`, that largely improves the state-of-the-art performance on a public benchmark for patient health prediction in intensive care units^9^. Due to its resource-efficient, automated design, our method is able to learn useful features from raw and heterogeneous input without laborious feature-engineering or impractical hardware demands. The model can deal with highly sparse, raw streaming inputs that have errors and missing values. As a result we improve patient health prediction over clinically used severity scores^21^ and modern, often much more complex and computationally expensive neural models^7,8^. Besides identifying important features that were used by others^9,22^, our approach uses important features that are intuitively sound upon closer inspection. Since (global) real world application in hospitals requires minimization of human labor, time, hardware and extensibility costs, we propose multiple modeling choices to conserve those resources, while also producing substantial improvements over state-of-the-art methods.

## Data Availability

The MIMIC III dataset is publicly available.

https://github.com/oguzserbetci/EffiCare

## Acknowledgments

This project was funded by the EU’s Horizon 2020 research and innovation program, grant No 780495 (BigMedilytics), as well as through the “Bundesministerium für Bildung und Forschung” projects CORA4NLP and XAINES.

## Footnotes

1. Project code: https://github.com/oguzserbetci/EffiCare. Paper to be published in amia.org/amia2020 via PubMed.

2. Originally published in 2017. Nature publication in 2019.

3. GeForce GTX 1080 Ti with 11GB memory, 10 Core CPU and 32GB RAM.

4. https://github.com/DFKI-NLP/MoRTy

5. https://github.com/copenlu/tx-ray

6. https://deepai.org/machine-learning-glossary-and-terms/disentangled-representation-learning

7. Due to space limitations, we do not show frequency adjusted relevance, though this is easy in practice.

## Notes

### Competing Interest Statement

The authors have declared no competing interest.

### Clinical Trial

This work uses part of the publicly available MIMIC III dataset. Hence, there were no trials.

### Funding Statement

This work was funded by both: the EU's Horizon 2020 research and innovation program, grant No 780495 (BigMedilytics) and the German Federal Ministry of Education and Research via the projects CORA4NLP and XAINES.

### Author Declarations

not applicable. Used publicly available dataset.

## References

1. E Choi, MT Bahadori, E Searles, C Coffey, M Thompson, J Bost, J Tejedor-Sojo, and J Sun. Multi-layer Representation Learning for Medical Concepts. In KDD. ACM, 2016.

2. E Choi, MT Bahadori, J Sun, J Kulas, A Schuetz, and W Stewart. Retain: An interpretable predictive model for healthcare using reverse time attention mechanism. In NeurIPS, pages 3504–3512, 2016.

3. Q Lu, Y Li, N de Silva, S Kafle, J Cao, D Dou, TH Nguyen, P Sen, B Hailpern, and B Reinwald. Learning electronic health records through hyperbolic embedding of medical ontologies. In BCB ‘19. ACM Press, 2019.

4. E Choi, MT Bahadori, L Song, WF Stewart, and J Sun. GRAM: graph-based attention model for healthcare representation learning. In Proc. of the 23rd ACM SIGKDD, 2017.

5. T Ma, C Xiao, and F Wang. Health-ATM: A deep architecture for multifaceted patient health record representation and risk prediction. In SIAM International Conference on Data Mining, 2018.

6. H Song, D Rajan, JJ Thiagarajan, and A Spanias. Attend and diagnose: Clinical time series analysis using attention models. In 32nd AAAI. AAAI press, 2018.

7. L Ma, C Zhang, Y Wang, W Ruan, J Wang, W Tang, X Ma, X Gao, and J Gao. ConCare: Personalized Clinical Feature Embedding via Capturing the Healthcare Context. In 34th AAAI. AAAI, 2019.

8. L Ma, J Gao, Y Wang, C Zhang, J Wang, W Ruan, W Tang, X Gao, and X Ma. AdaCare: Explainable Clinical Health Status Representation Learning via Scale-Adaptive Feature Extraction and Recalibration. In 34th AAAI. AAAI, 2019.

9. H Harutyunyan, H Khachatrian, DC Kale, G Ver Steeg, and A Galstyan. Multitask learning and benchmarking with clinical time series data. Scientific Data, 2017.

10. N Tomašev, X Glorot, JW Rae, M Zielinski, H Askham, A Saraiva, A Mottram, C Meyer, S Ravuri, I Protsyuk, et al. A clinically applicable approach to continuous prediction of future acute kidney injury. Nature, 2019.

11. P Bojanowski, E Grave, A Joulin, and T Mikolov. Enriching word vectors with subword information. TACL, 2017.

12. N Rethmeier and B Plank. Morty: Unsupervised learning of task-specialized word embeddings by autoencoding. In RepL4NLP@ACL, 2019.

13. S Gehrmann, H Strobelt, R Krüger, H Pfister, and A M. Rush. Visual interaction with deep learning models through collaborative semantic inference. IEEE TVCG, 2019.

14. N Rethmeier, VK Saxena, and I Augenstein. TX-Ray: Quantifying and Explaining Model-Knowledge Transfer in (Un-)Supervised NLP. In 36th UAI, 2020.

15. D Erhan, Y Bengio, A Courville, and P Vincent. Visualizing higher-layer features of a deep network. University of Montreal, 2009.

16. M Sundararajan, A Taly, and Q Yan. Axiomatic attribution for deep networks. In 34th ICML. JMLR, 2017.

17. AEW Johnson, TJ Pollard, L Shen, HL Li-wei, M Feng, M Ghassemi, B Moody, P Szolovits, LA Celi, and RG Mark. MIMIC-III, a freely accessible critical care database. Scientific data, 2016.

18. BC Kwon, M Choi, JT Kim, E Choi, Young B Kim, S Kwon, J Sun, and J Choo. RetainVis: Visual analytics with interpretable and interactive recurrent neural networks on electronic medical records. IEEE TVCG, 25, 2019.

19. T Pham, T Tran, D Phung, and S Venkatesh. DeepCare: A Deep Dynamic Memory Model for Predictive Medicine. In Advances in Knowledge Discovery and Data Mining. Springer International Publishing, 2016.

20. T Saito and M Rehmsmeier. The precision-recall plot is more informative than the ROC plot when evaluating binary classifiers on imbalanced datasets. PloS one, 2015.

21. JIF Salluh and M Soares. ICU severity of illness scores: APACHE, SAPS and MPM. Current opinion in critical care, 2014.

22. S Purushotham, C Meng, Z Che, and Y Liu. Benchmarking deep learning models on large healthcare datasets. Journal of Biomedical Informatics, 2017.

